# The cost of expanding Ethiopia’s salt iodization program to include multiple micronutrients

**DOI:** 10.1101/2025.04.16.25325958

**Authors:** Katherine P Adams, Dawd Gashu, Elias Asfaw Zegeye, Venkatesh Mannar, Levente L Diosady, N Ananth, E Louise Ander

## Abstract

**Background:** With near universal consumption of salt and technological advances that have made its fortification with multiple micronutrients feasible, salt has great potential for public health impact as a delivery vehicle for not only iodine but for multiple micronutrients. Decisions around modifying existing salt standards to include one or more additional micronutrients should consider not only potential impacts but also stakeholder-specific cost implications.

**Objective:** We aimed to estimate the total and incremental cost of expanding Ethiopia’s salt iodization program to include folic acid (dual fortified salt), folic acid and vitamin B12 (triple fortified salt), or folic acid, vitamin B12, and zinc (quadruple fortified salt).

**Methods:** We developed a set of activity– and ingredients-based cost models to estimate salt fortification costs over a 10-year time horizon. Cost model assumptions and parameters were primarily based on interviews with Ethiopian stakeholders in industry and government and non-governmental partners.

**Results:** Over the period 2024-2033, the estimated annual average cost of Ethiopia’s existing salt iodization program was ∼$2.1 million (2021 US dollars), or ∼$7/metric ton (MT) of fortified salt (∼$0.02/capita/year). Expanding the program to include folic acid would increase the annual average cost to ∼$2.5 million, or ∼$8.30/MT (∼$0.02/capita/year). Annually, the costs of triple and quadruple fortified salt programs were ∼$18 million (∼$59/MT; $0.13/capita) and $19 million (∼$63/MT; ∼$0.14/capita), respectively. Premix costs accounted for approximately half of the total cost of the iodized and dual fortified salt programs and ∼90% of triple and quadruple fortified salt. Industry and government costs represented smaller cost shares.

**Conclusions:** If Ethiopia considers modifying its existing salt iodization standard to include one or more additional micronutrient, there will be many important considerations, including costs and affordability. The cost estimates presented here can complement evidence on the potential for multiple fortified salt to improve dietary adequacy and reduce deficiency in Ethiopia.

## Introduction

Micronutrient deficiencies, sometimes referred to as hidden hunger, remain a significant public health problem globally (Bailey, West Jr & Black, 2015; Muthayya, Rah, Sugimoto et al., 2013; Victora, Christian, Vidaletti et al., 2021; Stevens, Beal, Mbuya et al., 2022). The consequences of micronutrient deficiencies can be severe, including adverse pregnancy outcomes (low birthweight, preterm birth, and neural tube defects, among others), impaired growth and cognitive development, decreased immune function, decreased work capacity, and mortality, all generating significant costs to individuals and societies (King, Brown, Gibson et al., 2016; Zimmermann, 2009; Black, 2008; Bailey et al., 2015).

The prevalence of micronutrient deficiencies tends to be higher among populations in low– and middle-income countries (LMICs) and with poor dietary diversity (Arimond, Wiesmann, Becquey et al., 2010). In Ethiopia, an estimated 72% of the population is zinc deficient based on serum zinc (Belay, Gashu, Joy et al., 2021). The majority of women (77.9%) have low RBC folate concentrations, consistent with increased risk of neural tube defect (NTD)-affected pregnancies (Sisay, Tamirat, Sandalinas et al., 2022). While there are no current estimates of the national rate of NTDs in Ethiopia, a systematic review of fifteen studies reported a pooled prevalence of 63.3 NTD cases per 10,000 children (Bitew, Worku, Alebel et al., 2020), a figure considerably higher than the worldwide prevalence of 18.6 per 10,000 live births (Blencowe, Kancherla, Moorthie et al., 2018). On the other hand, the prevalence of some micronutrient deficiencies are relatively low in Ethiopia, with 8% of women and adolescent girls and 15% of children deficient in iron and less than 10% of women and adolescent girls deficient in vitamin B12 (Ethiopian Public Health Institute, 2023). The economic cost of nutrient deficiencies and other forms of malnutrition in Ethiopia is high, estimated to cost Ethiopia 16.5% of its annual gross domestic product (Ethiopian Ministry of Health, 2013).

Globally, iodine deficiency has been largely addressed through the introduction of universal salt iodization programs. In 1994, the World Health Organization (WHO) recommended the iodization of salt to help eliminate iodine deficiency and associated health risks (World Health Organization, UNICEF & ICCIDD, 1994), and the current WHO guidelines identify universal salt iodization and sodium reduction strategies as compatible and cost-effective strategies to both reduce the global burden of non-communicable diseases and control iodine deficiency disorders (World Health Organization, 2022). As of 2020, salt iodization programs have been implemented on either a mandatory or voluntary basis in 145 countries, and 88% of the households worldwide consume salt that is fortified with iodine (Zimmermann & Andersson, 2021).

In the 1990s, Ethiopia introduced a universal salt iodization program, and although the program has been marked by periods of success with high coverage and periods of low coverage following times of conflict, currently ∼89% of households consume adequately iodized salt (Ethiopian Public Health Institute, 2018; Asfaw, Tamiru & Belachew, 2022; Yusufali, Frohmann, Chuko et al., 2022). More recently, Ethiopia introduced legislation mandating the fortification of wheat flour with zinc and B vitamins, including folic acid and vitamin B12 (Ethiopian Standards Agency, 2022). However, fortifiable wheat flour is consumed by only 30% of the population, mostly concentrated in urban areas (Ethiopian Public Health Institute, 2023). Adherence to iron-folic acid supplementation among pregnant women is also low (41.4%) (Sendeku, Azeze & Fenta, 2020), and contrary to the WHO recommendation to avert NTDs (World Health Organization, 2012a), a majority of women (71.5%) tend to start taking IFA supplements at later stage of their pregnancy (Gebremichael & Welesamuel, 2020). Further, given the links between periconceptual folate status and the risk of having an NTD-affected pregnancy (Molloy, Mills, Kirke et al., 1999), implementing strategies to improve folate levels among women before pregnancy is critical for NTD prevention (Martinez, Benavides-Lara, Arynchyna-Smith et al., 2023). Food or condiment fortification with folic acid has been identified as a feasible, effective strategy for improving maternal folate status and preventing NTDs, provided the program is well designed and implemented, including choosing a food vehicle that is consumed regularly by most of the population and adherence to fortification standards is monitored and enforced (Martinez et al., 2023; Tesfay, Hailu, Habtetsion et al., 2023).

Although salt is synonymous with iodine fortification, given its near universal consumption in fairly consistent and self-limiting quantities, the seasoning has the potential to also safely and effectively deliver other micronutrients (Matthias, McDonald, Archer et al., 2022). Double fortification of salt with iodine and iron has been introduced in India and reaches millions of consumers, primarily through social safety net programs (Diosady, Mannar & Krishnaswamy, 2019; Puri, Rekhi, Thomas et al., 2022; Moorthy & Rowe, 2021; Jadhav & Mannar, 2021). Technology to add folic acid, vitamin B12, and/or zinc in addition to iron has also been developed (Modupe & Diosady, 2021) and is being assessed for nutritional impact in India (McDonald, 2021; McDonald, Brown, Goh et al., 2022). Recent dietary modeling work showed the potential for multiple fortified salt to reduce the prevalence of zinc inadequacy by 6 to 51 percentage points and folate inadequacy by 2 to 56 percentage points among the Ethiopian population (Saje, Gashu, Joy et al., 2024), and a randomized trial to assess the nutritional impact and safety of salt fortified with iodine and folic acid is currently underway in Ethiopia (Brown, Tessema, McDonald et al., 2024). In terms of consumer acceptability, a recent study in Tanzania found that both double fortified and quadruple fortified salt were equally acceptable to consumers as standard iodized salt (Mdoe, Mannar, Paulo et al., 2023), and a recent acceptability trial in Ethiopia found that, despite slight color changes (from white to faint yellow), fine and coarse salt sprayed with potassium iodate and folic acid were acceptable to consumers in urban and rural Ethiopia (Tesfaye, Tessema, Arnold et al., 2025). Despite this growing body of evidence on the potential for salt to deliver multiple micronutrients to populations, including those that are often hard to reach with other micronutrient interventions, not a lot is known about the cost of multiply-fortified salt, especially outside the context of double fortified salt in India.

The Micronutrient Action Policy Support (MAPS) project is co-creating a web-hosted tool to estimate micronutrient deficiency risks and explore pathways to improve the micronutrient adequacy of diets (https://micronutrient.support/). The cost-effectiveness module of the tool enables users to estimate and compare the cost and cost-effectiveness of alternative micronutrient intervention programs. Using cost models we adapted from a salt iodization cost model that we developed for use in the MAPS tool, our objectives were to present estimates of the cost of Ethiopia’s salt iodization program, and the total and incremental cost of expanding Ethiopia’s salt fortification program to also include (1) folic acid (dual fortified salt), (2) folic acid and vitamin B12 (triple fortified salt), and (3) folic acid, vitamin B12, and zinc (quadruple fortified salt). These estimates will provide policymakers in Ethiopia with important evidence on the potential cost implications to different stakeholder groups of expanding the current salt iodization standard to provide one or more additional micronutrients.

## Methods

### Multiple fortification technology

Salt in Ethiopia is currently iodized at industrial-scale salt refining facilities, with salt typically iodized by spraying the salt with potassium iodate (Yusufali et al., 2022). Fortification of salt with iodine and folic acid can be accomplished by adding folic acid dissolved in sodium carbonate (to maintain alkaline conditions) to the potassium iodate solution and spraying the micronutrients together (Modupe & Diosady, 2021; Modupe, Siddiqui, Jonnalagadda et al., 2021; McGee, Sangakkara & Diosady, 2017). As such, salt refineries in Ethiopia could dual-fortify salt with iodine and folic acid without any additional equipment and with minimal changes to their current fortification processes. The addition of vitamin B12 and/or zinc along with folic acid is more complicated. Testing has shown that vitamin B12 is unstable when included in a spray solution containing folic acid (Modupe & Diosady, 2021), and iodine degrades quickly when exposed to zinc fortificants in a spray solution (Vatandoust, Krishnaswamy, Li et al., 2023). Salt fortification including folic acid and vitamin B12 and/or zinc is best accomplished when iodine is sprayed and the additional micronutrients are extruded and mixed with the iodized salt (Modupe & Diosady, 2021; Vatandoust et al., 2023).

The extrusion process involves first mixing the micronutrient premix with semolina flour, vegetable oil, and water and then extruding the dough in a cold forming extruder fitted with a very fine angel-hair pasta die. The extrudate is then cut to match the size of grains of salt then shaped using a spheronizer and dried. The extruded fortified salt-like grains (i.e., extruded premix) would then undergo color masking by spraying the extruded particles with a color masking agent (such as titanium dioxide) and then encapsulated by coating the particles with hydroxypropyl methylcellulose (HPMC) and soy stearin (Vatandoust et al., 2023). With technology transferred from the University of Toronto, JVS Foods Pvt Ltd in Jaipur, India has the capacity to produce, at large scales, extruded premix for salt fortified with iodine and iron (Jadhav, Mannar & Wesley, 2019; Diosady, Mannar & Menon, 2018). JVS also has the capacity to produce extruded multiple micronutrient premix (Diosady et al., 2018; McDonald, 2021). Given this, we assumed that for triple and quadruple fortified salt, the extruded premix would be produced at the JVS facility, imported into Ethiopia, and distributed to salt refineries. Regardless of where the extruded premix is produced, to support triple or quadruple salt fortification in Ethiopia, refineries would require the installation of blending equipment (including a ribbon or screw blender) to mix the extruded premix with iodized salt to produce multiple fortified salt.

### Cost models

We developed a set of activity– and ingredients-based cost models to estimate the economic cost of the current salt iodization program in Ethiopia and the hypothetical cost of expanding that program to mandate dual (iodine and folic acid), triple (iodine, folic acid, and vitamin B12), or quadruple (iodine, folic acid, vitamin B12, and zinc) fortified salt. The cost models, which were developed in Microsoft Excel, were structured based on the set of activities required to plan, execute, and manage each alternative salt fortification program. For each activity, we identified a series of inputs, or “ingredients”, that go into performing the activity (e.g., labor, supplies, etc.) and populated each activity in the model with the estimated number of units and associated unit cost of each input needed to undertake the activity.

Costs were estimated over a 10-year time horizon (2024-2033), which allowed for two years of planning and initiation activities associated with expanding Ethiopia’s salt iodization program to include additional micronutrients. Costs were defined from a societal perspective, meaning all costs were included regardless of who incurs them. As such, cost estimates accounted for costs that might be paid by the salt industry (refineries), the Ethiopian government, and development partners, as well as costs potentially passed on to salt consumers. Costs were expressed in 2021 US dollars (USD). For input costs reported in USD, where necessary we adjusted the value to 2021 USD using the Bureau of Economic Analysis implicit price deflators for gross domestic product (Bureau of Economic Analysis, 2020). For input costs reported in Ethiopian Birr, we first adjusted to the 2021 value using the local consumer price index (World Development Indicators, 2024) and then converted to USD using the average 2021 exchange rate.

### Cost model assumptions and parameter values

The model developed to estimate the cost of Ethiopia’s current salt iodization program was comprised strictly of recurring costs in each year of the 10-year time horizon. These recurring costs included the cost of potassium iodate, including shipping and local storage and transportation (note that Ethiopia does not tax micronutrient premix, so no taxes or import duties were included). Recurring costs also included industry-related costs to fortify the salt (including labor, power/fuel, annualized equipment costs, and maintenance costs), costs to conduct internal quality assurance and quality control (QA/QC) activities at refineries (internal quantitative iodine tests via titration and WYD spectrophotometer, external lab tests, etc.), and overhead/administration. Finally, they included government regulatory and monitoring costs (refinery inspections and monitoring, import, commercial, and household monitoring), training/retraining of government personnel, social marketing and advocacy, and overhead/administration.

Each model developed to estimate the cost of expanding the salt fortification program to include multiple micronutrients also included two years of expansion costs to account for costs associated with government planning for and revision of the national salt fortification standards, the acquisition of new refinery/fortification facility equipment needed to blend the extruded premix with iodized salt (relevant for triple and quadruple fortified salt), the acquisition of new equipment needed for monitoring, and training for industry personnel involved in salt fortification and training/capacity building for government personnel involved in monitoring of the salt fortification program. We assumed that salt iodization would continue as usual in years one and two and that salt fortification with multiple micronutrients would begin in year three and continue through the end of the modeling time horizon. For the dual fortified salt scenario, activities contributing to recurring costs were very similar to those for iodized salt except the additional cost of folic acid for spraying onto salt alongside potassium iodate as well as some additional monitoring costs to test for folic acid in salt. For the triple and quadruple fortified salt scenarios, recurring costs also accounted for the additional cost of the extruded premix (including shipping and local storage and handling), additional labor, power/fuel, equipment, and equipment maintenance cost, and additional management/overhead. Because we assumed recurring government monitoring activities would be focused on testing for iodine and folic acid, government monitoring costs were the same for dual, triple, and quadruple fortified salt.

For all scenarios, underlying model assumptions and parameter values, including unit costs and quantities, were based on primary data collection, local and international expert input, and secondary data sources. We conducted interviews with salt refineries in Ethiopia to understand the activities required to iodize salt and conduct QA/QC and to generate estimates of the associated personnel and supply costs. We also conducted interviews with government personnel involved in monitoring of the salt iodization program to accurately characterize current monitoring and enforcement activities and to generate estimates of the cost of undertaking those activities. Some model assumptions and parameter values were informed by secondary data, including published standards for salt iodization in Ethiopia, United Nations World Population Prospects for national population projections over the 10-year time horizon (United Nations, 2019), and published and grey literature. Equipment costs were annualized using a 3% discount rate and assuming a useful life of 10 years.

For dual, triple, and quadruple fortified salt, we modeled two different potential fortification levels (**Table 1**). The first set of fortification levels (scenario 1) were based on current salt consumption patterns (7.4 grams/day among women of reproductive age) estimated from urinary sodium excretion in the National STEPS survey (Challa, Tadesse, Mudie et al., 2017), and assuming 90% of salt intake is from discretionary intake and manufactured food items (Saje et al., 2024). The second set of fortification levels (scenario 2) were based on a hypothetical scenario in which discretionary salt consumption in Ethiopia decreased to ∼5 grams/day, in line with WHO recommendations to limit sodium intake to < 2000 mg/d (World Health Organization, 2012b). Except for iodine, in both scenarios the fortification levels were developed based on modeling of dietary data in Addis Ababa and the Somali Region (regions known for lowest and highest micronutrient deficiency, respectively, compared to other regions), with the fortification levels for each salt consumption level selected to maximize micronutrient intake and also ensure risk of high intake remained below 5% (Saje et al., 2024). For scenario 1, we used Ethiopia’s current salt iodization standard (30 mg/kg) while for scenario 2 we assumed salt would be iodized to 39 mg/kg, in line with WHO guidelines when salt consumption is ∼5 gram/day among adults (World Health Organization, 2014). Note that cost estimates did not account for costs associated with sodium reduction campaigns or the cost of revising fortification standards in response to reduced salt consumption levels. Given the low prevalence of iron deficiency in Ethiopia (estimated to be 8% among WRA and adolescent girls and 13% among children 6-59 months) (Ethiopian Public Health Institute, 2023), we did not model the inclusion of iron. The prevalence of vitamin B12 deficiency is also low (<10% among WRA and adolescent girls) (Ethiopian Public Health Institute, 2023), but we included B12 because it plays an important role in folate metabolism and could therefore improve the effectiveness of fortifying salt with folic acid (Scott, 1999).

**Table 1.**
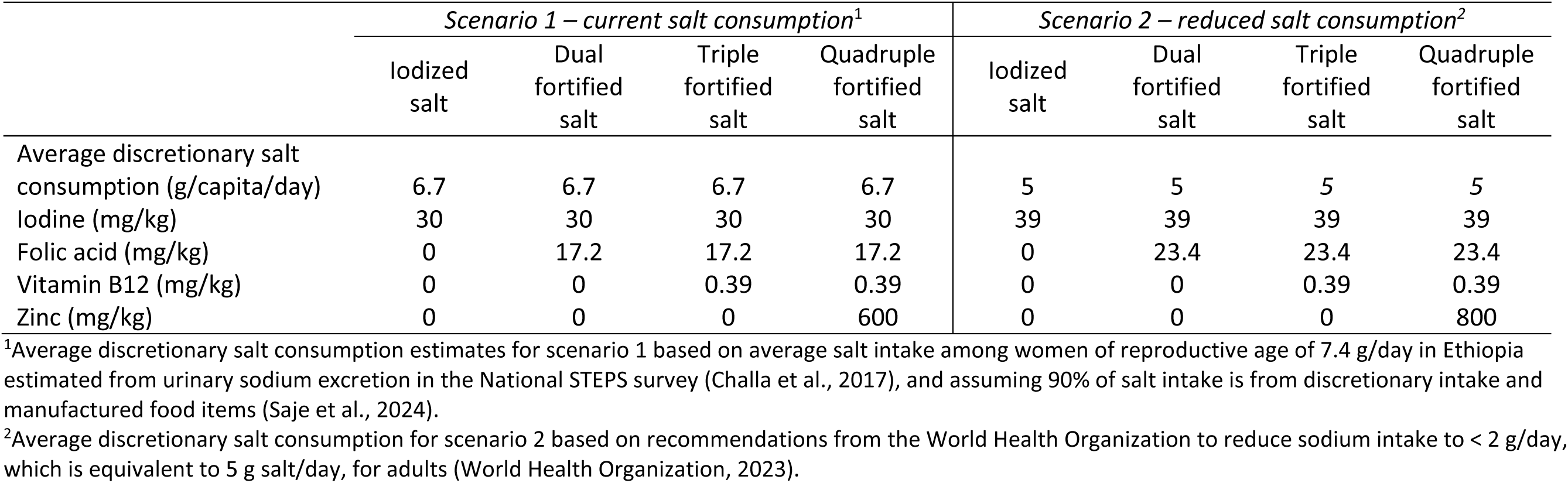
Salt fortification cost modeling scenarios.

For sprayed micronutrients (potassium iodate for each type of fortified salt and folic acid for dual fortified salt), we estimated premix costs using a premix cost calculator that was developed to enable estimation of the cost of micronutrient premix for fortified salt and other food vehicles with the flexibility to adjust fortification levels, micronutrient fortificants, and fortificant prices. Fortificant prices in the premix cost calculator were informed by estimates provided by an international large-scale food fortification (LSFF) program expert in November 2021. Because the triple and quadruple salt fortification scenarios involved extruded and encapsulated micronutrients that we assumed would be produced in India and imported into Ethiopia, the cost estimates of the extruded premixes were based on estimates provided by JVS Foods Pvt Limited as informed by the company’s experience producing premixes for efficacy trials in India in 2021 and validated by market cost estimates obtained from Indiamart (www.indiamart.com). As JVS Foods Pvt Limited is currently the only private premix manufacturer with the technology and capacity to produce extruded fortified premix for multiple fortified salt, these estimates represent the best available source of cost data. However, in recognition of uncertainty around these cost estimates, we conducted sensitivity analyses around these and other uncertain parameters (described below). Recent estimates of industry compliance with Ethiopia’s salt fortification standard suggest that most (89%) of the salt is fortified with iodine (Ethiopian Public Health Institute (EPHI) & ICF, 2016). As such, we modeled costs assuming (1) all discretionary salt in the food system in Ethiopia is fortifiable, and (b) 89% of discretionary salt and salt used in manufactured food items is fortified to the national standard.

### Sensitivity analyses

Because multiple fortified salt is a hypothetical micronutrient intervention in Ethiopia, some of our cost model parameter values were based on cost data from other contexts as well as informed assumptions about how a multiple salt fortification program in Ethiopia would be designed and implemented. To assess the influence of uncertain parameter values on the total and incremental cost of expanding Ethiopia’s salt iodization program to include additional micronutrients, for the current salt consumption patterns scenario (scenario 1), we conducted several sets of sensitivity analyses around particularly uncertain parameters.

First, because micronutrient premix is often the most expensive component of a LSFF program and because, apart from salt fortified with iodine and iron, multiple fortified salt has not been produced for use at scale and our cost estimates are based on cost data from one company, we varied the price of the micronutrient premix up and down by 30% (including the cost of potassium iodate). We also recognize uncertainty around the additional costs salt refineries would face to fortify salt with multiple micronutrients, so we also varied up and down by 30% the total cost of salt refinery fortification costs, which includes labor, power/fuel, annualized equipment costs, and equipment maintenance costs. There is also uncertainty in how the government of Ethiopia would monitor a multiple fortified salt program, so we assessed the impact of this uncertainty by varying total government monitoring costs (including refinery monitoring, salt import monitoring, and household monitoring) up and down by 30%. Finally, we estimated best-case and worst-case scenarios by simultaneously decreasing the price/cost of each of these uncertain parameters by 30% (the best-case scenario) and simultaneously increasing their price/cost by 30% (the worst-case scenario).

## Results

### Primary results

We estimated that, over the 10-year time horizon of 2024-2033, the annual average cost of Ethiopia’s current salt iodization program was ∼$2.1 million (2021 USD), equating to ∼$7 per metric ton (MT) of fortified salt and ∼$0.02 per capita per year (**Table 2**, scenario 1). Expanding the current program to include folic acid was estimated to increase the annual average cost by ∼$415,000 to approximately $2.5 million and to increase the average cost per MT of fortified salt to ∼$8.30 (note that the cost per capita rounds to ∼$0.02 in both cases, although it is slightly higher for dual fortified salt compared to iodized salt). For triple fortified salt, which involves mixing iodized salt with extruded premix that includes folic acid and vitamin B12 fortificants, we estimated that the annual average cost would increase to ∼$18 million. Compared to the current iodization program, triple fortified salt was estimated to increase the cost of fortification by ∼$52/MT of salt to ∼$59/MT, and increase the annual average cost per capita to $0.13, or ∼$0.11 higher than the cost per capita of Ethiopia’s current salt iodization program. Finally, the estimated annual average cost of quadruple fortified salt was slightly over $19 million, or ∼$63 per MT and ∼$0.14 per capita per year.

**Table 2.** Estimated total and incremental cost of salt fortification in Ethiopia based on current salt consumption.

|  | Iodized salt | Dual fortified salt | Triple fortified salt | Quadruple fortified salt |
| --- | --- | --- | --- | --- |
| **Scenario 1 – current salt consumption** |  |  |  |  |
| **Total cost<sup>1</sup>** |  |  |  |  |
| Annual average total cost | \$2,115,000 | \$2,531,000 | \$18,009,000 | \$19,014,000 |
| Annual average cost per capita<sup>2</sup> | \$0.02 | \$0.02 | \$0.13 | \$0.14 |
| Annual average premix cost<sup>3</sup> per MT of fortified salt | \$2.7 | \$3.7 | \$43.1 | \$45.7 |
| Annual average total cost per MT of fortified salt | \$7.0 | \$8.3 | \$59.3 | \$62.6 |
| **Incremental cost<sup>4</sup>** |  |  |  |  |
| Annual average total cost | - | \$416,000 | \$15,894,000 | \$16,899,000 |
| Annual average cost per capita<sup>1</sup> | - | \$0.003 | \$0.11 | \$0.12 |
| Annual average premix cost<sup>2</sup> per MT of fortified salt | - | \$1.0 | \$40 | \$43 |
| Annual average total cost per MT of fortified salt | - | \$1.4 | \$52 | \$56 |
| **Scenario 2 – reduced salt consumption** |  |  |  |  |
| **Total cost<sup>1</sup>** |  |  |  |  |
| Annual average total cost | \$1,987,000 | \$2,410,000 | \$17,474,000 | \$18,318,000 |
| Annual average cost per capita<sup>2</sup> | \$0.01 | \$0.02 | \$0.12 | \$0.13 |
| Annual average premix cost<sup>3</sup> per MT of fortified salt | \$3.5 | \$4.9 | \$55.9 | \$58.9 |
| Annual average total cost per MT of fortified salt | \$8.7 | \$10.6 | \$76.6 | \$80.3 |
| **Incremental cost<sup>4</sup>** |  |  |  |  |
| Annual average total cost | - | \$423,000 | \$15,487,000 | \$16,331,000 |
| Annual average cost per capita<sup>1</sup> | - | \$0.003 | \$0.11 | \$0.12 |
| Annual average premix cost<sup>2</sup> per MT of fortified salt | - | \$1.4 | \$52 | \$55 |
| Annual average total cost per MT of fortified salt | - | \$1.9 | \$68 | \$72 |

kg, kilogram; MT, metric ton.
<sup>1</sup>Costs modeled over 10-year time horizon (2024-2033) and reported in undiscounted 2021 US dollars. Total costs are rounded to the nearest thousand and costs per MT are rounded to the nearest tenth.
<sup>2</sup>Ethiopia population estimates based on the 2019 World Population Prospects total population estimates, 2024-2033 (United Nations, 2019).
<sup>3</sup>Premix costs include the cost of micronutrient fortificants, and, in the case of triple and quadruple fortified salt, the cost of additional ingredients and the production of extruded fortified salt-like grains.
<sup>4</sup>Incremental costs calculated as the difference in cost between iodized salt and the alternative multiple fortified salt scenario.

Based on the hypothetical scenario in which average salt consumption in Ethiopia fell to WHO recommended levels of 5 gram/day among adults (with modeled fortification levels correspondingly increased), the estimated annual average costs of iodized, dual, triple, and quadruple fortified salt would be similar to annual average cost estimates based on current salt consumption patterns (**Table 2**, scenario 2). However, given that a smaller quantity of fortified salt would be required to meet demand, the average price per MT of fortified salt would be higher. Specifically, the estimated cost per metric ton of fortified salt was ∼$8.70 for iodized salt, ∼$10.60 for dual fortified salt (iodine and folic acid), ∼$76.60 for triple fortified salt (iodine, folic acid and B12), and ∼$80/MT for quadruple fortified salt (iodine, folic acid, B12 and zinc). The corresponding annual average cost per capita estimates were $0.01, $0.02, $0.12, and $0.13, respectively.

Activity-specific cost estimates and proportional contributions of each activity to total annual average costs based on current salt consumption patterns (scenario 1) are reported in **Table 3**. Recurring premix costs for iodized and dual fortified salt made up approximately half of the total intervention program costs (47% and 55%, respectively). This was followed by (1) recurring industry-related fortification costs, including labor, power/fuel, annualized equipment costs, and equipment maintenance costs, representing 26% of the total salt iodization program costs and 22% of the total cost of double fortified salt, and (2) recurring refinery management, administration, and overhead costs (16% and 14%, respectively). For the triple and quadruple fortified salt programs, recurring premix costs accounted for over 90% of the total annual average cost. Note that the micronutrient fortificants contributed 24% and 54% to the total cost of the extruded, encapsulated premix for triple and quadruple fortified salt, respectively, while other ingredients and the manufacturing process accounted for the remained of the premix costs. In all cases, other start-up and recurring costs made up a small proportion (6% or less) of the total cost of the programs. In terms of the potential distribution of costs across stakeholder groups, refinery-related costs (excluding premix) accounted for ∼49% of the annual average cost of the iodized salt program and ∼41% of the total cost of the dual fortified salt program. Given that premix costs are significantly higher for triple and quadruple fortified salt, these refinery-related costs represented ∼8% of the total cost of these fortification programs. For each potential salt fortification program, average annual government-related costs were <5% of the total annual average cost.

**Table 3.** Activity-specific cost and share of total cost estimates of salt fortification in Ethiopia based on current salt consumption.

|  | *Scenario 1 – current salt consumption* |  |  |  |  |  |  |  |
| --- | --- | --- | --- | --- | --- | --- | --- | --- |
|  | Iodized salt |  | Dual fortified salt |  | Triple fortified salt |  | Quadruple fortified salt |  |
|  | Average annual cost (2021 USD) | Percent of total cost | Average annual cost (2021 USD) | Percent of total cost | Average annual cost (2021 USD) | Percent of total cost | Average annual cost (2021 USD) | Percent of total cost |
| **Start-up/scale-up costs** |  |  |  |  |  |  |  |  |
| Relabeling, annualized cost | \$0 | 0% | \$1,000 | 0% | \$1,000 | 0% | \$1,000 | 0% |
| Training for salt refinery personnel, annualized cost | \$0 | 0% | \$1,000 | 0% | \$1,000 | 0% | \$1,000 | 0% |
| Government planning (reformulation of standards, adoption/reformulation of M&E plan, etc.), annualized cost | \$0 | 0% | \$2,000 | 0% | \$3,000 | 0% | \$3,000 | 0% |
| Training/capacity building for government M&E personnel, annualized cost | \$0 | 0% | \$2,000 | 0% | \$2,000 | 0% | \$2,000 | 0% |
| **Recurring costs** |  |  |  |  |  |  |  |  |
| Premix, including shipping and taxes | \$1,002,000 | 47% | \$1,388,000 | 55% | \$16,432,000 | 91% | \$17,437,000 | 92% |
| Salt refinery fortification costs, including labor, power/fuel, annualized equipment costs, and maintenance costs | \$546,000 | 26% | \$546,000 | 22% | \$835,000 | 5% | \$835,000 | 4% |
| Salt refinery fortification QA/QC activities | \$125,000 | 6% | \$125,000 | 5% | \$125,000 | 1% | \$125,000 | 1% |
| Salt refinery internal training/retraining | \$22,000 | 1% | \$22,000 | 1% | \$22,000 | 0% | \$22,000 | 0% |
| Salt refinery management, administration, and overhead related to fortification | \$347,000 | 16% | \$347,000 | 14% | \$491,000 | 3% | \$491,000 | 3% |
| Government inspections and monitoring of salt refineries | \$8,000 | 0% | \$13,000 | 1% | \$13,000 | 0% | \$13,000 | 0% |
| Government monitoring of imported salt | \$2,000 | 0% | \$3,000 | 0% | \$3,000 | 0% | \$3,000 | 0% |
| Government monitoring of salt at markets and other retail outlets | \$2,000 | 0% | \$12,000 | 0% | \$12,000 | 0% | \$12,000 | 0% |
| Government household monitoring | \$14,000 | 1% | \$14,000 | 1% | \$14,000 | 0% | \$14,000 | 0% |
| Social marketing/advocacy | \$9,000 | 0% | \$9,000 | 0% | \$9,000 | 0% | \$9,000 | 0% |
| Capacity building/training for food control agency personnel/monitors/lab technicians | \$14,000 | 1% | \$14,000 | 1% | \$14,000 | 0% | \$14,000 | 0% |
| Government management, administration, and overhead | \$24,000 | 1% | \$32,000 | 1% | \$32,000 | 0% | \$32,000 | 0% |
| Total costs |  |  |  |  |  |  |  |  |
| Annual average premix cost<sup>1</sup> | \$1,002,000 | 47% | \$1,388,000 | 55% | \$16,432,000 | 91% | \$17,437,000 | 92% |
| Refinery-related annual average total cost | \$1,040,000 | 49% | \$1,042,000 | 41% | \$1,474,000 | 8% | \$1,474,000 | 8% |
| Government-related annual average total cost | \$73,000 | 3% | \$101,000 | 4% | \$103,000 | 1% | \$103,000 | 1% |
| Total annual average cost | \$2,115,000 | 100% | \$2,531,000 | 100% | \$18,009,000 | 100% | \$19,014,000 | 100% |

M&E, monitoring and evaluation; QA/QC, quality assurance/quality control
Costs modeled over 10-year time horizon (2024-2033) and reported in undiscounted 2021 US dollars rounded to the nearest thousand.
<sup>1</sup>Premix costs include the cost of micronutrient fortificants, and, in the case of triple and quadruple fortified salt, the cost of additional ingredients and the production of extruded fortified salt-like grains.

The breakdown of activity-specific costs, their contributions to total costs, and the possible distribution of costs across stakeholder groups based on the reduced salt consumption scenario are presented in **Table 4**. For each possible salt fortification program, these estimates were very similar to the current salt consumption scenario, with micronutrient premix representing ∼50% of the total annual average cost of the iodized and dual salt fortification programs and over 90% for triple and quadruple fortified salt.

**Table 4.**
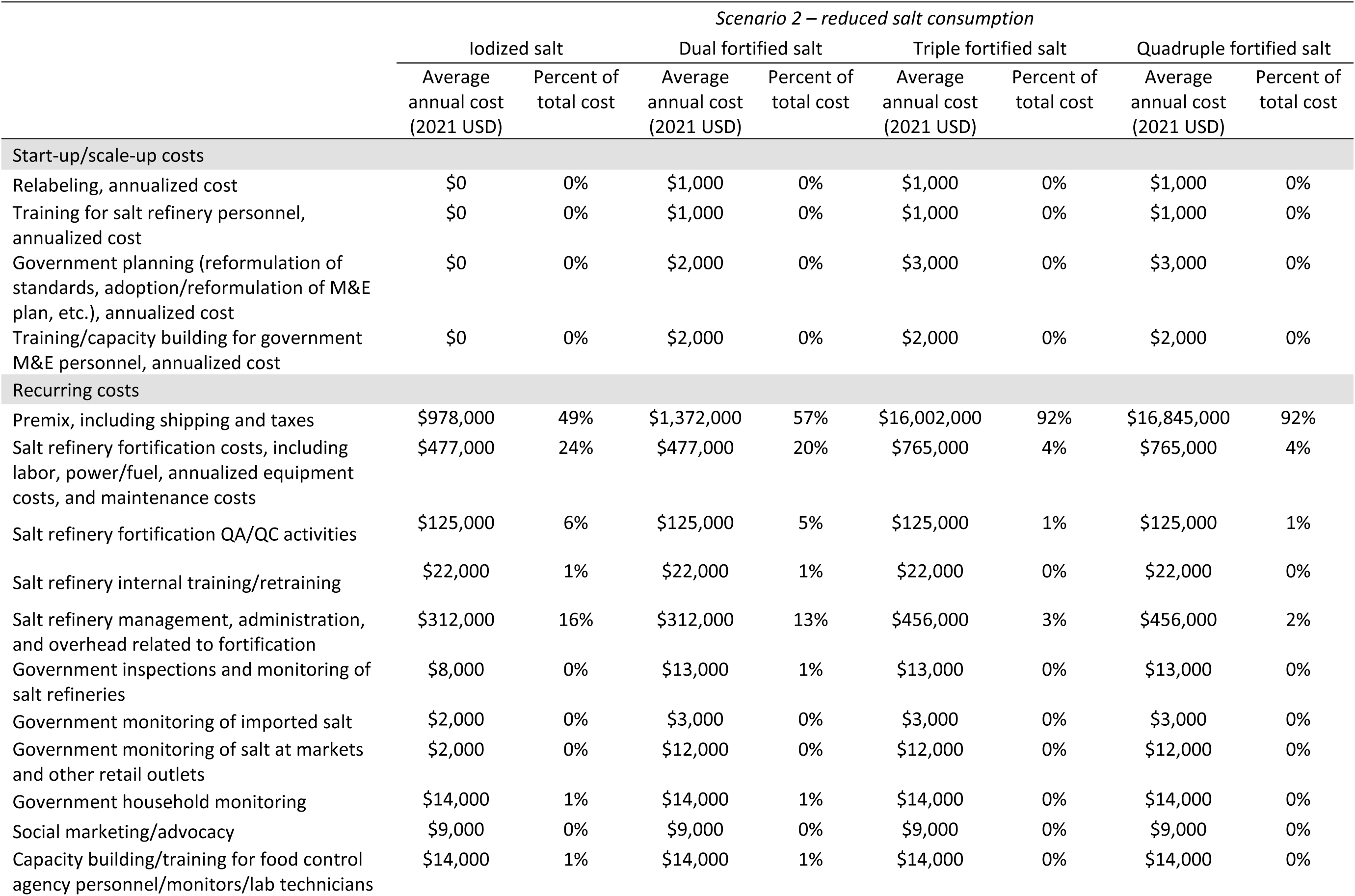
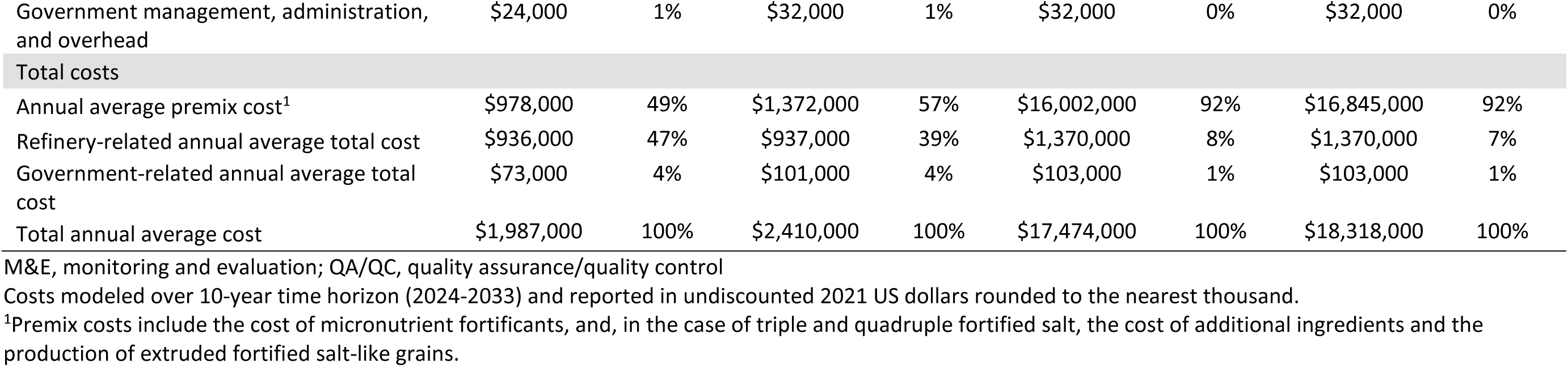
Activity-specific cost and share of total cost estimates of salt fortification in Ethiopia based on reduced salt consumption.

### Sensitivity analysis

Variation in the estimated annual average cost per MT of fortified salt across several sources of uncertainty are presented in **Figure 1**. For each type of fortified salt, varying the price of the micronutrient premix (and the extruded premix in the case of triple and quadruple fortified salt) up and down by 30% had the largest impact on the annual average cost per MT of fortified salt, potentially increasing or decreasing the annual average cost of iodized salt by ∼$1.00/MT, the cost of double fortified salt by ± ∼$1.40/MT, and the cost of triple and quadruple salt by ± ∼$16-$18/MT. Thirty percent variation in refinery fortification-related costs increased or decreased the cost per MT by ∼$1 for dual, triple, and quadruple fortified salt, while 30% variation in government monitoring costs had a small impact (∼$0.05) on the estimated annual average cost per MT. With all sources of cost variation considered simultaneously, the “worst-case” estimated annual average cost was $10.55/MT for dual fortified salt, $77.85/MT for triple fortified salt, and $82.15 for quadruple fortified salt. At the other extreme, the “best case” estimates of the annual average cost per MT were $6.11, $41.22, and $43.53 for dual, triple, and quadruple fortified salt, respectively.

**Figure 1.**
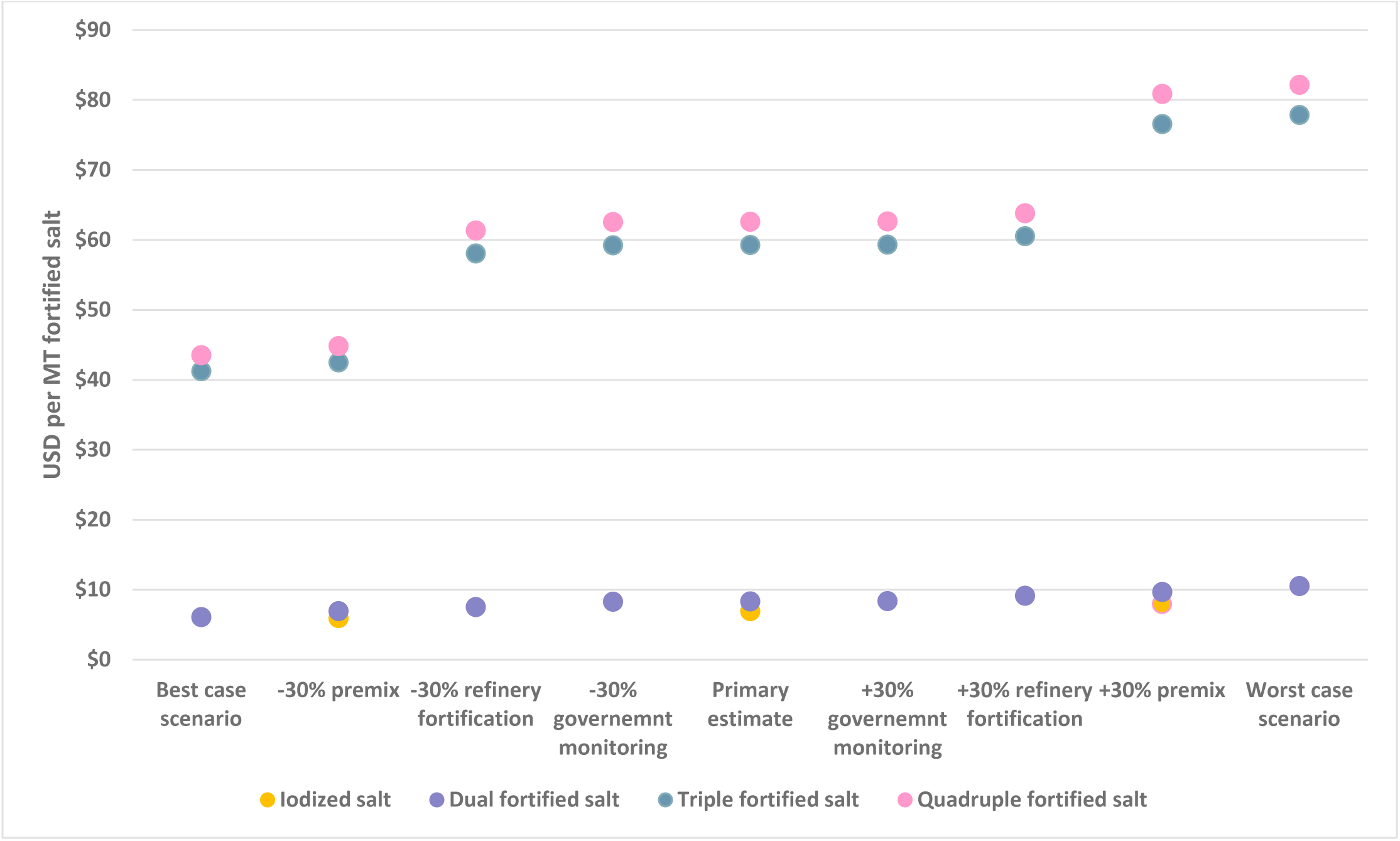
Sensitivity analysis – estimated annual average cost per metric ton (MT) of fortified salt based on current salt consumption patterns (scenario 1) with variation in key cost model inputs.

## Discussion

Universal salt iodization has played a fundamental role in the global reduction of iodine deficiency disorders, with substantial health and economic benefits (Gorstein, Bagriansky, Pearce et al., 2020; Zimmermann, 2023). Expanding salt iodization programs to fortify salt with multiple micronutrients has the potential to build on this success and improve the adequacy of multiple micronutrients in diets. However, decisions around expanding salt iodization programs should be evidence-based. Beyond salt fortified with iodine and iron, the body of evidence on the potential impacts of salt fortified with other micronutrients is growing. An upcoming randomized controlled trial (RCT) in the Oromia Region of Ethiopia will provide evidence on the potential impact of salt fortified with iodine and folic acid on the folate status of women of reproductive age (ClinicalTrials.gov, 2024; Brown et al., 2024). In Punjab, India, a RCT will provide evidence on the impact of quintuply-fortified salt (iodine, iron, folic acid, vitamin B12, and zinc) on the biomarker status of women of reproductive age and preschool-age children (McDonald et al., 2022). Finally, recent modeling work using dietary recall data showed that multiple-fortified salt could lead to substantial reductions in the prevalence of inadequate zinc and folate intakes in both urban and rural Ethiopia (Saje et al., 2024). In this paper, based on detailed activity-based cost models, we provide complementary evidence on the potential cost of using salt as a food vehicle to enhance the micronutrient adequacy of diets in Ethiopia beyond iodine.

We found that, from a societal costing perspective and based on current salt consumption patterns, the dual fortified salt program (iodine and folic acid), which could rely on existing salt iodization technology, would increase the annual average cost of the salt fortification program from ∼$7/MT of fortified salt to ∼$8.30/MT, corresponding to an annual average increase in the cost per capita of less than $0.01. The preferred technology for triple (iodine, folic acid, and vitamin B12) or quadruple (iodine, folic acid, vitamin B12, and zinc) fortified salt requires extruding the micronutrient fortificants into a salt-like grain that is then encapsulated and mixed with iodized salt (Modupe & Diosady, 2021; Vatandoust et al., 2023). We estimated that this extrusion and encapsulation process, in addition to the added cost of the additional micronutrient fortificants, additional refinery fortification-related costs, and additional government monitoring and evaluation costs would increase the total annual average cost of the program substantially, from just over ∼$2 million per year to ∼$18 and $19 million annually, translating to $0.11 and $0.12 higher estimated annual average costs per capita. Note that these additional costs would not only deliver one or two additional micronutrients but would also likely improve the effectiveness of fortifying salt with folic acid because of the role of vitamin B12 in folate metabolism (Scott, 1999). Under a hypothetical scenario in which salt consumption in Ethiopia dropped to an average of 5 grams/day among adults, necessitating higher fortification levels to help meet requirements but also a smaller total quantity of salt in the food system to fortify, the estimated total and incremental costs were similar to those based on current salt consumption patterns.

The price of iodized salt in Addis Ababa in October, 2024 was ∼$0.27 per kg for a relatively inexpensive salt packaged in a plastic bag (although the market price varies depending on type of salt, packaging, etc.). Relative to this, the incremental cost estimates represent a ∼0.5% increase in the price for dual fortified salt, a ∼19% increase for triple fortified salt, and a ∼21% increase in the price of quadruple fortified salt. While it is unlikely that all of these incremental costs would be passed on to salt consumers, even if just premix costs were passed on to consumers, this would still represent a sizeable increase in the price of salt in the case of triple and quadruple fortified salt. At the same time, it is important to acknowledge the potential additional health benefits that may be conferred to the population if salt also delivered vitamin B12 (to aid in folate metabolism) and/or zinc to deficient populations in Ethiopia. Issues such as affordability and the tradeoffs between higher salt prices and public health benefits would need to be considered by decision-makers in Ethiopia. Moreover, although the incremental cost of triple and quadruple fortified salt is low in absolute terms at $0.11-$0.12 per capita per year, the cost relative to iodized and dual fortified salt may be a barrier to adoption in Ethiopia and other LMICs. Future research in this space to identify alternative technologies for multiple fortified salt at a lower cost could help reduce this potential barrier.

The cost estimates presented in this study should be interpreted with several limitations in mind. First, because a multiple fortified salt program in Ethiopia is hypothetical, some of our cost inputs and assumptions about how the program might operate were based on experiences in other contexts and on informed assumptions. We conducted sensitivity analyses to assess the impact of some of this uncertainty, but actual costs incurred in practice may still vary from our estimates. Also, our cost estimates reflected current industry compliance with the national salt iodization standard. If compliance with a revised standard that required fortifying salt with multiple micronutrients was lower (or higher) than current compliance, this would impact the cost of the program. Related, studies assessing the feasibility of multiple fortified salt have been conducted with dried refined salt (Modupe & Diosady, 2021; Modupe et al., 2021; McGee et al., 2017; Vatandoust et al., 2023). While most of Ethiopia’s salt supply for human consumption is based on production processes that produce high-quality, refined salt, some proportion (< 30%) is produced using practices that results in lower-quality salt (Yusufali et al., 2022). As such, the technical feasibility of fortification of lower-quality salt with multiple micronutrients would need to be considered. Another limitation is that our cost estimates for triple and quadruple fortified salt are based on the assumption that the extruded premix would be produced in India and imported into Ethiopia. The Ethiopian market is currently too small to justify a local premix facility, but if the market for encapsulated micronutrient fortification expanded and Ethiopia developed capacity for local production of the extruded premix, this may change program costs. Finally, because spraying salt with folic acid has been shown to turn the salt yellow, consumer acceptability of dual fortified salt may require education and advocacy efforts (Modupe et al., 2021). In India, organoleptic changes to salt as a result of the inclusion of iron have negatively impacted consumer acceptance and uptake, thus curtailing demand for double fortified salt in the open market (Moorthy & Rowe, 2021). Although recent evidence from Ethiopia on the acceptability of dual and triple fortified salt suggests consumer acceptability will not be a significant barrier in that context (Tesfaye et al., 2025), this may still be an important consideration, particularly if organoleptic changes are more pronounced with, e.g., the fortification of lower-quality salt.

Study strengths include the development of detailed, 10-year activity-based cost models, the modeling of several different multiple fortified salt alternatives, and modeling costs based on not only current salt consumption patterns but also a scenario in which salt consumption in Ethiopia decreased to a level closer to the WHO levels recommended for sodium reduction. Sodium reduction efforts need not conflict with salt fortification programs (World Health Organization, 2014). The modeling results based on discretionary salt consumption of ∼5 gram/day demonstrate how reductions in salt consumption in response to, e.g., sodium reduction campaigns, can be integrated into salt fortification programs by adjusting fortification levels to ensure dietary requirements are still met while also understanding the potential cost implications of adjusting fortification standards.

Salt, with its nearly universal consumption across all population groups and recent technological advances that have made its fortification with multiple micronutrients feasible and reliable (i.e., with micronutrient stability and retention) (Modupe & Diosady, 2021), has great potential for public health impact as a delivery vehicle for multiple micronutrients. If Ethiopia considers modifying its existing salt iodization standard to include one or more additional micronutrient, there will be many important considerations, including which micronutrients to include and at what fortification levels, consumer acceptability of the chosen formulation, how the program would be regulated and enforced, feasibility to adhere with revised standards for smaller-scale salt refineries, etc. Another important consideration will be how the cost of the salt fortification program would likely change, which stakeholder groups would be called upon to pay those higher costs, and whether those higher costs would be affordable to each stakeholder group. This paper provides evidence on the potential incremental costs of expanding Ethiopia’s current salt iodization program to include folic acid, folic acid and vitamin B12, or folic acid, vitamin B12, and zinc. This evidence can complement evidence on the potential impact of multiple fortified salt. To further build this evidence base, next steps could include estimating the cost-effectiveness and/or cost-benefit of multiply-fortified salt in Ethiopia.

## Funding

This work was supported, in whole or in part, by the Gates Foundation [INV-002855]. The conclusions and opinions expressed in this work are those of the author(s) alone and shall not be attributed to the Foundation. Under the grant conditions of the Foundation, a Creative Commons Attribution 4.0 License has already been assigned to the Author Accepted Manuscript version that might arise from this submission. Please note works submitted as a preprint have not undergone a peer review process.

## Data Availability

The Excel cost model to estimate the cost of salt iodization is publicly and freely available without restriction at https://doi.org/10.5281/zenodo.14564050. Excel cost models to estimate the cost of dual, triple, and quadruple fortified salt will be made available upon request from the corresponding author.

## Acknowledgements

The authors gratefully acknowledge the stakeholders in industry and the government in Ethiopia who participated in interviews and provided costing data. We would also like to acknowledge the Micronutrient Intervention Modeling Project for sharing previous Ethiopia cost modeling work.

## Contributions

KPA, DG, and ELA designed the study. KAP, DG, EAZ, VM, and NA collected the data. LLD advised on the appropriate multiple fortified salt technology for each scenario. KPA developed the cost models and wrote the first draft of the manuscript. All authors contributed to the data interpretation and revisions of the manuscript, and read and approved the final manuscript.

## Competing interests

All authors have no competing interests to declare.

## Notes

### Competing Interest Statement

The authors have declared no competing interest.

